# Migration and Outbreaks of Vaccine-Preventable Disease in Europe: A Systematic Review

**DOI:** 10.1101/2021.01.08.21249473

**Authors:** Anna Deal, Rachael Halliday, Alison Crawshaw, Sally Hayward, Amelia Burnard, Kieran Rustage, Jessica Carter, Anushka Mehrotra, Felicity Knights, Ines Campos-Matos, Azeem Majeed, Jon S. Friedland, Michael Edelstein, Sandra Mounier-Jack, Sally Hargreaves, in collaboration with the ESCMID Study Group for Infections in Travellers and Migrants (ESGITM)

## Abstract

**Background:** Migrant populations (defined as foreign-born) are one of several under-immunised groups in the EU/EEA, yet little is known about how they are affected by outbreaks of vaccine-preventable diseases (VPDs). This information is vital to develop targeted strategies to improve the health of diverse migrant communities and to assess risk factors and correlations with major European peaks in incidence of key VPDs over time.

**Methods:** We did a systematic review (PROSPERO CRD42019157473; Medline, EMBASE, and Global Health January 2000 to October 2019) adhering to PRISMA guidelines, to identify studies on VPD outbreaks (measles, mumps, rubella, diphtheria, pertussis, polio, hepatitis A, *N meningitidis*, and *H influenzae*) in migrants residing in the EU/EEA and Switzerland.

**Results:** 45 studies were included, reporting on 47 distinct VPD outbreaks across 13 countries (26 [55%] were reported between 2010 and 2020, including 16 [34%] since 2015). Most reported outbreaks involving migrants were of measles (n=24; 6578 total cases), followed by varicella (n=11; 596 cases), hepatitis A (n=7; 1510 cases), rubella (n=3; 487 cases) and mumps (n=2; 295 cases). 19 (40%) of outbreaks, predominantly varicella and measles, were reported in temporary camps or shelters for asylum seekers and refugees. Of 11 varicella outbreaks, 82% were associated with adult migrants. Half of measles outbreaks (n=12) were associated with migrants from Eastern European countries, often involving migrants of Roma ethnicity.

**Conclusions:** Migrants represent one of several under-immunised groups involved in VPD outbreaks in Europe, with adult and child refugees and asylum seekers residing in shelters or temporary camps at particular risk, alongside specific nationality groups. Vulnerability varies by disease, setting, and individual demographics, highlighting the importance of tailoring strategies for implementing catch-up vaccination to specific groups, alongside the strengthening of routine data collection, in order to meet regional and global vaccination targets. Better understanding vaccine uptake and demand issues in migrant groups, and reducing the barriers they face to accessing vaccination services, is urgently needed, with direct implications for COVID-19 vaccine delivery at the current time. Strengthening vaccine delivery to migrant populations will require a greater focus on co-designing vaccine uptake strategies in close collaboration with affected communities.

**Funder:** *NIHR*

## Introduction

Migration to the EU/EEA and Switzerland has risen substantially in recent years and includes a diverse group of asylum seekers, refugees, labour migrants, EU/EEA internal migrants, and undocumented migrants from many regions of the world, with considerable implications for health services (1). Migrants are considered to be one of several under-immunised groups in Europe (1-3). The European Vaccine Plan asks Member States to pay particular attention to migrants, international travellers, and other marginalised communities to ensure their access to effective and culturally appropriate immunisation services (4). Migrants in Europe may come from countries with limited or disrupted health and vaccination systems and may therefore be unaligned with the vaccination schedule of their host country. Certain settings, such as refugee camps and immigration detention centres, may also be extremely conducive to vaccine-preventable disease (VPD) outbreaks with unsanitary living conditions, lack of access to healthcare often combined with low vaccination coverage of residents (5).

Migrants may face barriers in access to health services, exclusion from services due to lack of entitlement to free health care, and inconsistencies in delivery of care, including for catch-up vaccinations and other preventative health-care services (7, 8). There are currently shortfalls in catch-up vaccination delivery for migrants on arrival to the EU/EEA, particularly among older age-groups outside routine childhood vaccination schedules. Language is often a key barrier for migrants accessing vaccination services (6) as well as a lack of systems in place for catch-up vaccination in adolescent and adult migrants (7). Some migrants may also fear approaching healthcare providers, including vaccination services, because of links with immigration authorities (8). Data suggest that low confidence in vaccination could also contribute to low immunisation coverage of some migrant groups (9, 10), yet it is unclear to what extent migrants – or specific nationality groups – are vaccine hesitant (11). These factors hold immediate relevance to current COVID-19 vaccination roll-out, with ethnic minority groups in one study associated with low intent to vaccinate for COVID-19 in the UK and US (12).

Although migrant communities have been affected by VPD outbreaks in EU/EEA countries, the link between migration and outbreaks has been poorly elucidated to date. It is essential that further research is carried out to identify drivers and risk-factors to inform targeted strategies for improving vaccination uptake in at-risk populations. Several major measles outbreaks have been recorded in Europe in recent years, with over 60,000 cases reported since January 2015 (data to September 2020), thought to involve under-vaccinated adult migrants (13, 14). Major measles clusters in Greece, Belgium and Germany during 2019 suggested that migrants from Eastern European countries may be heavily affected compared to other populations (15-17). However, regional databases do not routinely collate data on VPD incidence disaggregated by migrant status, meaning the vaccination of migrants is currently a poorly evidenced aspect of public health policy in Europe. What data do exist is often not disaggregated and therefore gives little information on high-risk groups, including specific nationalities. These data are vital if we are to develop effective and targeted vaccination interventions to ensure immunisation coverage reaches established Herd Immunity Threshold (HIT) targets to prevent VPD outbreaks (18, 19). Ensuring high levels of vaccination coverage is a key priority for European countries through the European Vaccine Action Plan, to prevent outbreaks with unnecessary morbidity, mortality and costs to health care systems.

We therefore did a systematic review to collate data from all outbreaks of key VPDs involving migrants in EU/EEA countries and Switzerland, to identify key risk factors and drivers involved.

## Methods

### Search strategy and inclusion/exclusion criteria

We did a systematic literature review in line with Preferred Reporting Items for Systematic Reviews and Meta-Analyses (PRISMA) guidelines (20) and registered on PROSPERO (CRD42019157473). Primary outcomes were migrant cases of vaccine-preventable diseases as part of an outbreak; secondary outcomes were risk factors associated with having a VPD (age, nationality, migrant status, setting, immunisation status). We searched Embase, Global Health, and MEDLINE for peer-reviewed primary research reporting on VPD outbreaks involving migrant populations in the EU/EEA and Switzerland between January 1^st^ 2000 and May 22^nd^ 2020, with no language restrictions. Following a search of relevant systematic reviews and consulting with experts in the field, a Boolean search strategy was developed containing terms relating to migration, outbreaks of VPD and European location (see Supplementary file for full search terms).

We defined a migrant as any individual born outside of the country in which data were collected. We included papers reporting primary data from observational studies (e.g. cross-sectional, case-control or cohort studies and outbreak or case reports), conference abstracts and systematic reviews (where they presented data not published elsewhere). Comments, editorials, literature reviews and letters were excluded. Non-English papers were included to ensure that the review is representative of outbreaks occurring across Europe. Studies were eligible for inclusion if they reported primary data on one or more outbreaks involving at least one migrant in the EU/EEA and Switzerland where migrant status was clear.

We used the WHO definition to define an outbreak: “the occurrence of cases of disease in excess of what would normally be expected in a defined community, geographical area or season. An outbreak may occur in a restricted geographical area, or may extend over several countries. It may last for a few days or weeks, or for several years. A single case of a communicable disease long absent from a population, or caused by an agent (e.g. bacterium or virus) not previously recognized in that community or area, or the emergence of a previously unknown disease, may also constitute an outbreak and should be reported and investigated”(21). Therefore, for all diseases except polio, reports of single, unlinked cases of disease were excluded. Polio has been absent from the European region since 2002 (22, 23), therefore a single case would be considered an outbreak and included. Data not disaggregated into distinct outbreaks were included in the narrative synthesis but excluded from spatial mapping. For studies reporting on multiple outbreaks, data was extracted separately for each outbreak. Where multiple studies reported on the same outbreak, the most complete description of the outbreak was included, with multiple reports included when necessary to provide required data.

Outbreaks pertaining to the following diseases were included: diphtheria, *Haemophilus influenzae* type b (Hib), hepatitis A, measles, mumps, *N. meningitidis*, pertussis, polio and rubella. We searched for studies from the following EU/EEA countries and Switzerland: Austria, Belgium, Bulgaria, Cyprus, Croatia, Czech Republic, Denmark, Estonia, Finland, France, Germany, Greece, Hungary, Iceland, Ireland, Italy, Latvia, Liechtenstein, Lithuania, Luxembourg, Malta, the Netherlands, Norway, Poland, Portugal, Romania, Slovakia, Slovenia, Spain, Sweden, Switzerland, and the UK.

Exclusion criteria were studies in which data were not transparently reported for migrants or migrant status could not be identified, studies reporting on fewer than two cases of a disease (except for polio), on an expected number of cases for the area or on multiple but clearly unlinked cases. Studies reporting on cases of vaccine-derived polio as opposed to wild-type polio were also excluded.

### Data screening, extraction, and synthesis

AD carried out title and abstract screening, full text screening, data extraction, and quality assessment, with all steps duplicated by an independent second reviewer (SH, RH or AB), in line with PRISMA guidelines. Where decisions could not be reached, two further reviewers (SEH, KR) arbitrated. All titles and abstracts were screened for their relevance and eligibility. Full text screening was carried out for all potentially eligible studies, and reasons for exclusion recorded.

Data were extracted on the following, where available: Time period of outbreak (year, month, duration), location (country, setting i.e. temporary camps [defined as: refugee camps, shelters, asylum residences or detainment centres] or general community, population affected (migrant status [e.g. refugee, asylum seeker, students, workers], age-group, gender, nationality, ethnicity) number of cases, migrant proportion of cases, index case, reported immune status of cases (oral vaccine history or measured antibody titres), diagnostic methods and genotyping data. Where an outbreak resulted in cases in multiple settings, the setting where the majority of cases were found was reported.

Quality assessment of the included studies was carried out independently by two reviewers using the Joanna Briggs Institute (JBI) critical appraisal tool (20). 10 points were allocated to each study, with scores of 8-10 considered high quality; 5-7 medium, 1-4 low. Studies were not excluded based on quality assessment in order to increase transparency.

### Spatial mapping analysis

Using the ggplot2 package in R (version 3.6.1), the number of cases in all published outbreaks involving migrants was plotted against the GPS co-ordinates of the reported outbreak location. Where an exact location was not provided, the GPS co-ordinates of the country were used. The outbreaks were colour coded by disease type.

## Results

### Overview of included studies

45 studies were included, reporting 47 distinct VPD outbreaks across 13 EU/EEA countries and Switzerland (Figure 1). Twenty-one outbreaks involving migrants (45%) were reported in these countries between 2000 and 2010 and 26 (55%) were reported between 2010 and 2020, including 16 (34%) since 2015. Two reports, from Spain and Germany, did not disaggregate cases into distinct outbreaks, but presented all outbreak-related cases across a specific time period (24, 25). Measles had the highest number of reports of outbreaks involving migrants (n=24; 6496 cases), followed by varicella (n=11; 505 cases), hepatitis A (n=7; 1356 cases), rubella (n=3; 487 cases) and mumps (n = 2; 293 cases). Table 1 summarises the characteristics of all included studies. In terms of quality, 23 papers were of high quality, 16 medium and 6 low, with research not excluded on the basis of quality.

**Table 1.** Details of all included studies including study quality (based on JBI tool, 10-point scale: 8-10 high, 5-7 medium, 1-4 low)

| Author | Publication Year | Number of Outbreaks | Outbreak start Year | Disease | Country | Location | Cases | Setting | Quality |
| --- | --- | --- | --- | --- | --- | --- | --- | --- | --- |
| Boletin Epidemiológico de Madrid (44) | 2005 | 1 | 2005 | Rubella | Spain | Madrid | 460 | Community | 9 |
| Boletin Epidemiológico de Madrid (45) | 2007 | 1 | 2006 | Measles | Spain | Madrid | 174 | Community | 9 |
| Chaud (30) | 2017 | 1 | 2016 | Varicella | France | Calais | 351 | Camp | 8 |
| Ciancotti (25) | 2013 | multiple | 2003-2012 | Hepatitis A | Spain | Valencia | 152 | Community | 9 |
| Ciccullo (40) | 2018 | 1 | 2016 | Hepatitis A | Italy | Rome | 141 | Community | 9 |
| Curtale (46) | 2010 | 1 | 2006 | Measles | Italy | Lazio | 449 | Community | 7 |
| De Valliere (31) | 2011 | 3 | 2007, 2008 | Varicella | Switzerland | Vaud | 2, 2, 15 | Camp | 7 |
| Dominguez (48) | 2008 | 1 | 2006 | Measles | Spain | Catalonia | 381 | Community | 9 |
| Epiemiologisches bulletin (32) | 2010 | 1 | 2010 | Varicella | Germany | Munich | 7 | Camp | 8 |
| Filia (47) | 2006 | (see Curtale) |  |  |  |  |  |  | 5 |
| Gabutti (33) | 2003 | 1 | 2002 | Varicella | Italy | Salento | 60 | Camp | 9 |
| Garcia (63) | 2007 | 1 | 2005 | Mumps | Spain | Almeria | 145 | Community | 8 |
| Gee (49) | 2010 | 1 | 2009 | Measles | Ireland | Southern Ireland | 320 | Community | 4 |
| Georgakopoulou (50) | 2006 | 1 | 2005 | Measles | Greece | Northern Greece | 171 | Community | 7 |
| Georgakopoulou (51) | 2018 | 1 | 2017 | Measles | Greece | Southern Greece | 3150 | Community | 7 |
| Gold (52) | 2010 | 1 | 2010 | Measles | Germany | Munich | 48 | Community | 8 |
| Grammens (17) | 2017 | 1 | 2016 | Measles | Belgium | Wallonia | 177 | Community | 8 |
| Ivic Hofman (53) | 2011 | 1 | 2011 | Measles, Varicella | Croatia | Salvonski-Brod | 4, 10 | Community | 6 |
| Jones (26) | 2016 | 1 | 2016 | Measles | France | Calais | 13 | Camp | 8 |
| **Kieran (41)** | 2011 | 1 | 2010 | Hepatitis A | Ireland | Cork | 11 | Community | 3 |
| **Kuehne (24)** | 2016 | multiple | 2004-2014 | Hepatitis A, Measles, Mumps, Varicella | Germany | countrywide | 2, 82, 2, 91 | Camp | 7 |
| **Lampl (16)** | 2019 | 1 | 2018 | Measles | Germany | Regensburg | 11 | Camp | 8 |
| **Lemos (64)** | 2004 | 1 | 2002 | Rubella | Spain | Madrid | 19 | Community | 7 |
| **Lesens (27)** | 2016 | 1 | 2015 | Varicella | France | Calais | 12 | Camp | 9 |
| **Mancuso (59)** | 2008 | 1 | 2004 | Measles | Germany | Wiesbaden | 5 | Other | 7 |
| **Mankertz 2011 (62)** | 2011 | 2 | 2010 | Measles | Germany, Austria | Karlsruhe, Vienna, | 4, 6 | Camp, Community | 7 |
| **Martinez-Torres (65)** | 2009 | (see Boletin Epi de Madrid 2005) |  |  |  |  |  |  | 4 |
| **Melidou (55)** | 2012 | 1 | 2010 | Measles | Greece | countrywide | 126 | Community | 4 |
| **Mellou (28)** | 2017 | 1 | 2016 | Hepatitis A | Greece | countrywide | 188 | Camp | 9 |
| **Mellou (42)** | 2020 | 1 | 2017 | Hepatitis A | Greece |  | 294 | Community | 7 |
| **Michaelis (34)** | 2017 | 1 | 2015 | Hepatitis A | Germany | countrywide | 699 | Camp | 7 |
| **Nic Lochlainn (38)** | 2016 | 1 | 2014 | Measles | UK | London | 10 | Community | 7 |
| **Nordbo (39)** | 2016 | 1 | 2015 | Mumps | Norway | Trondheim | 148 | Community | 3 |
| **Perez Sautu (43)** | 2011 | 2 | 2006 | Hepatitis A | Spain | Catalonia | 3, 20 | Community | 3 |
| **Pervanidou (54)** | 2010 | (See Melidou) |  |  |  |  |  |  | 6 |
| **Roggendorf (67)** | 2012 | 1 | 2010 | Measles | Germany | Essen | 11 | Community | 8 |
| **Stefanoff (35)** | 2005 | 3 | 2003 | Measles | Poland | Czerwonym Borze, Lublinie, Białymstoku | 3, 13, 14 | Camp | 6 |
| **Takla (29)** | 2012 | 1 | 2010 | Measles | Germany | Neumunster | 8 | Camp | 9 |
| **Torner (61)** | 2008 | 1 | 2005 | Rubella | Spain | Barcelona | 8 | Community | 8 |
| **Vainio (57)** | 2011 | 1 | 2011 | Measles | Norway | Oslo | 10 | Community | 9 |
| **Vairo (89)** | 2017 | 1 | 2015 | Varicella | Italy | Latium | 41 | Camp | 9 |
| **Valdarchi (60)** | 2008 | 1 | 2005 | Varicella | Italy | Roma | 5 | Other | 10 |
| **Vierucci (58)** | 2010 | 1 | 2008 | Measles | Italy | Pisa | 44 | Community | 6 |
| **Werber (37)** | 2017 | 1 | 2014 | Measles | Germany | Berlin | 1344 | Camp | 9 |
| **Zhang (66)** | 2020 | 1 | 2017 | Varicella | UK | England | 4 | Camp | 8 |

**Figure 1.**
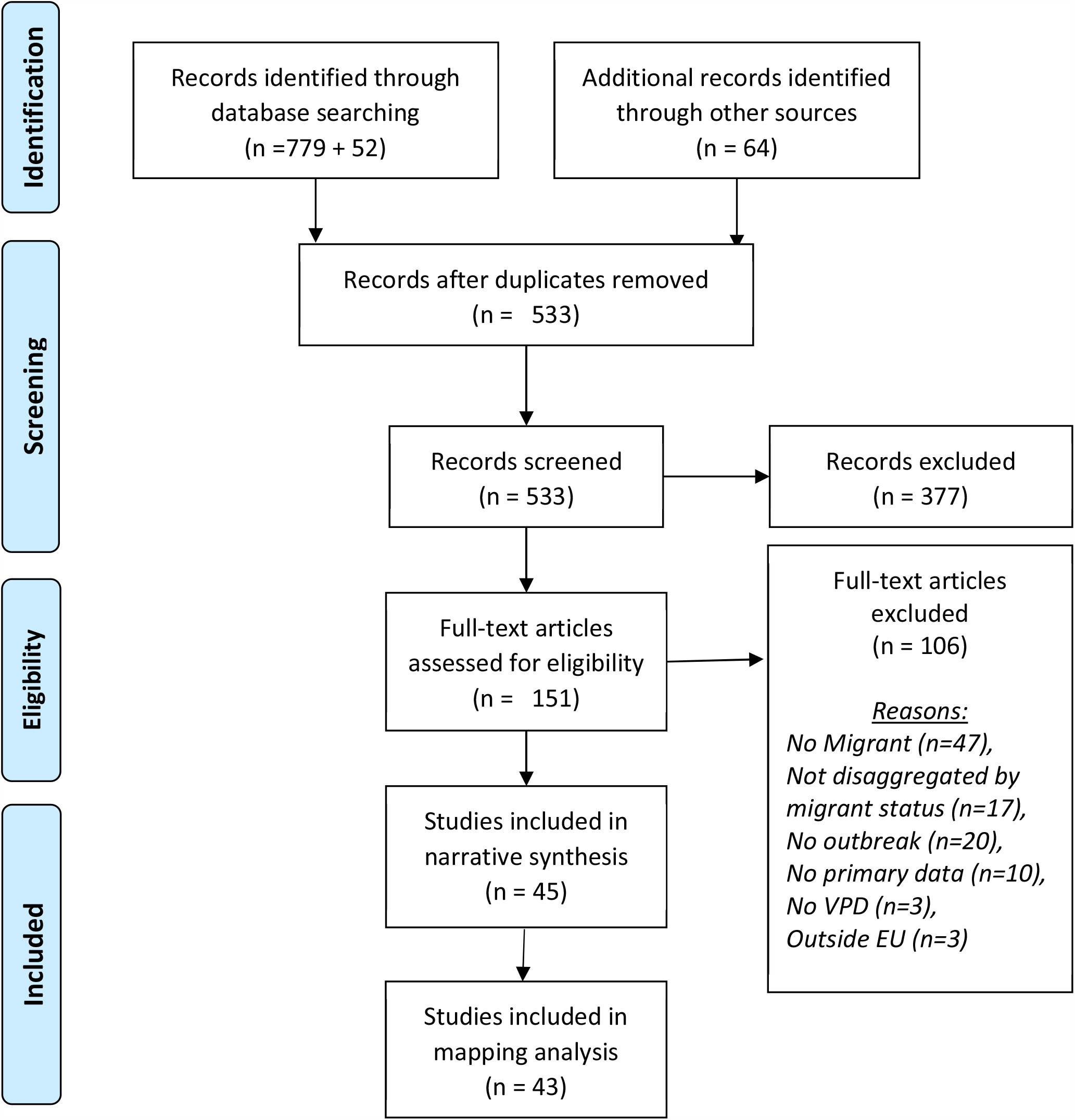
PRISMA diagram of included and excluded studies

### Outbreak Characteristics and Settings

In 35 (74%) included outbreaks, the index case was a migrant; of which 19 outbreaks (56%) only went on to subsequently infect migrants, and 6 (18%) only clearly reported migrant status for the index case. 19 outbreaks (40%) were reported from hosting facilities for refugees and asylum seekers (temporary camps, detention centres or mass accommodations) (16, 26-37), whereas 26 (55%) outbreaks involving migrants were reported in the general community (17, 25, 37-58). Other settings in which outbreaks were reported included an American military base (59) and a prison (60).

Of outbreaks occurring in hosting facilities for refugees and asylum seekers (n = 19), four reported spread to members of the local population. During these four outbreaks, the secondary cases among the local population were mostly limited; one case of hepatitis A in a health care worker (HCW) based in an affected camp (34), three HCWs and one volunteer infected with measles during an outbreak in the Calais refugee camps (26) and the son of an asylum centre worker infected with varicella (33). However, one outbreak of measles originating in asylum seekers (146 asylum seekers affected) in Berlin spread widely among the host population, resulting in 1,344 cases overall, 345 hospitalisations and the death of a 1-year old child (37). The most frequent outbreaks occurring in camp-settings were varicella (n=9) (24, 27, 30-33, 36) and measles (n=8) (16, 24, 26, 29, 35, 37), with nearly all (75%) varicella outbreaks occurring in these settings, as shown in Figure 2.

**Figure 2.**
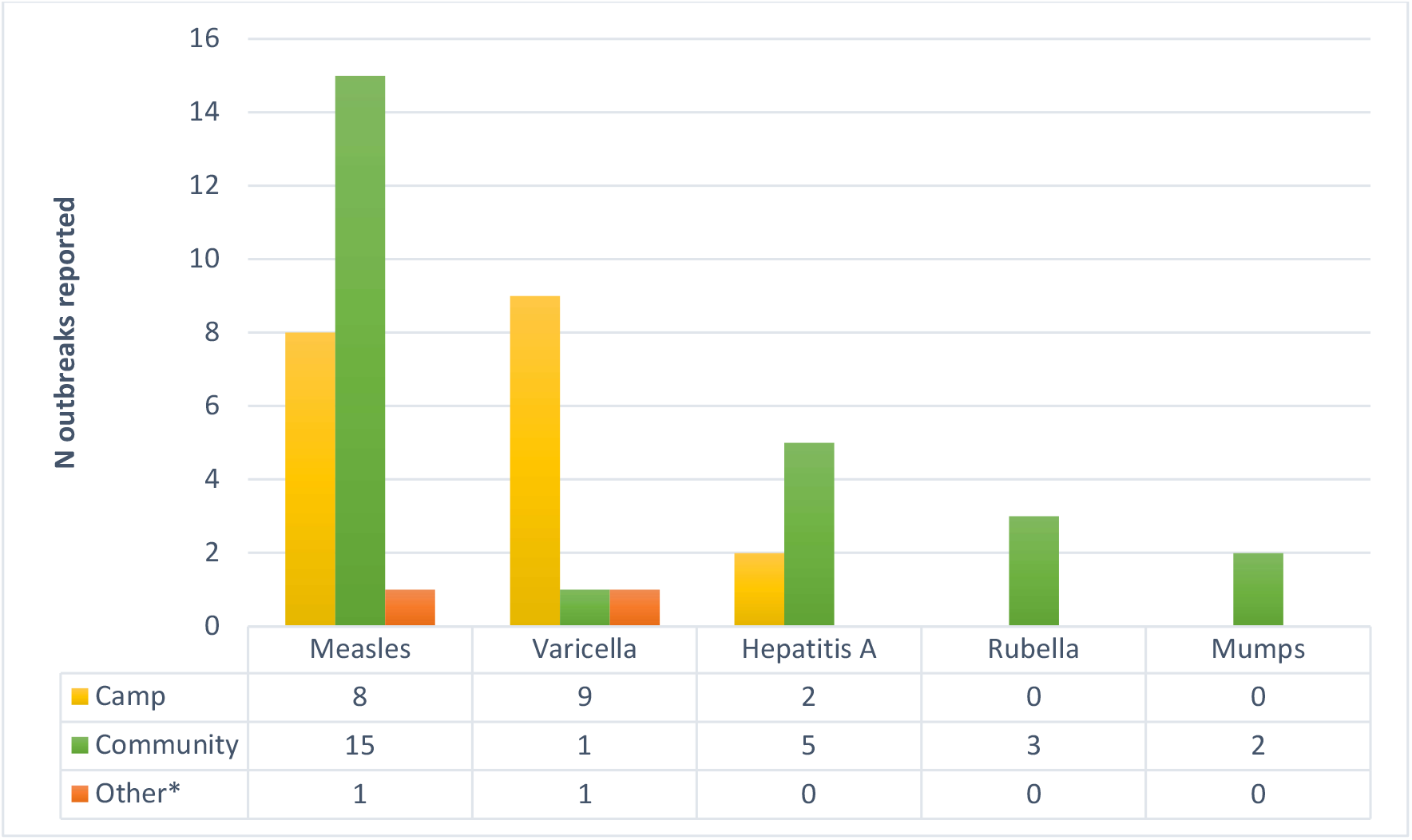
Settings of outbreaks involving migrants. *Other settings were a prison and a US military base.

Of outbreaks occurring in the community (n = 26), 17 (65%) reported a migrant index case. These included two small outbreaks of hepatitis A in which the index cases were internationally adopted children (41, 43), a rubella outbreak originating from a Brazilian woman living in Spain (61), a small measles outbreak with a Somali child as the index case (57) and multiple measles outbreaks with index cases from Eastern European countries, predominantly of Roma ethnicity. The most common diseases causing outbreaks in the general community were measles (n=15), hepatitis A (n=5) and rubella (n=3).

### Geographical distribution of VPDs: Spatial mapping analysis

Germany (n=9) (16, 29, 32, 34, 37, 52, 56, 59, 62), Spain (n=8) (43-45, 48, 61, 63-65), Italy (n=6) (33, 36, 40, 46, 47, 58, 60) and Greece (n=5) (28, 42, 50, 51, 54, 55) were the countries that most frequently reported outbreaks involving migrants between 2000 and 2020. Two thirds (n=6) of included outbreaks in Germany affected refugee or asylum-seeking populations, mostly in temporary camps, 78% (n=7) were measles outbreaks and all but one were reported in 2010 or later. All included outbreaks in Spain took place in the general community between 2000 and 2007. In Italy, the outbreaks included were equally distributed between 2000 and 2020 and half of outbreaks were of varicella. Outbreaks reported from Greece were predominantly measles (n=3) and all but one occurred in 2010 or later.

Other countries reporting outbreaks were Austria (n=1) (62), Belgium (n=1) (17), Croatia (n=2) (53), England (n=2) (38, 66), France (n=3) (26, 27, 30), Ireland (n=2) (41, 49), Norway (n=2) (39, 57), Poland (n=3) (35) and Switzerland (n=3) (31). Some clustering of diseases by country was observed, for example, rubella outbreaks involving migrants between 2000 and 2020 were only reported in Spain (61, 64, 65). Figure 3 highlights the case-load of VPD outbreaks (n=43) involving migrants by reported location, highlighting clustering.

**Figure 3.**
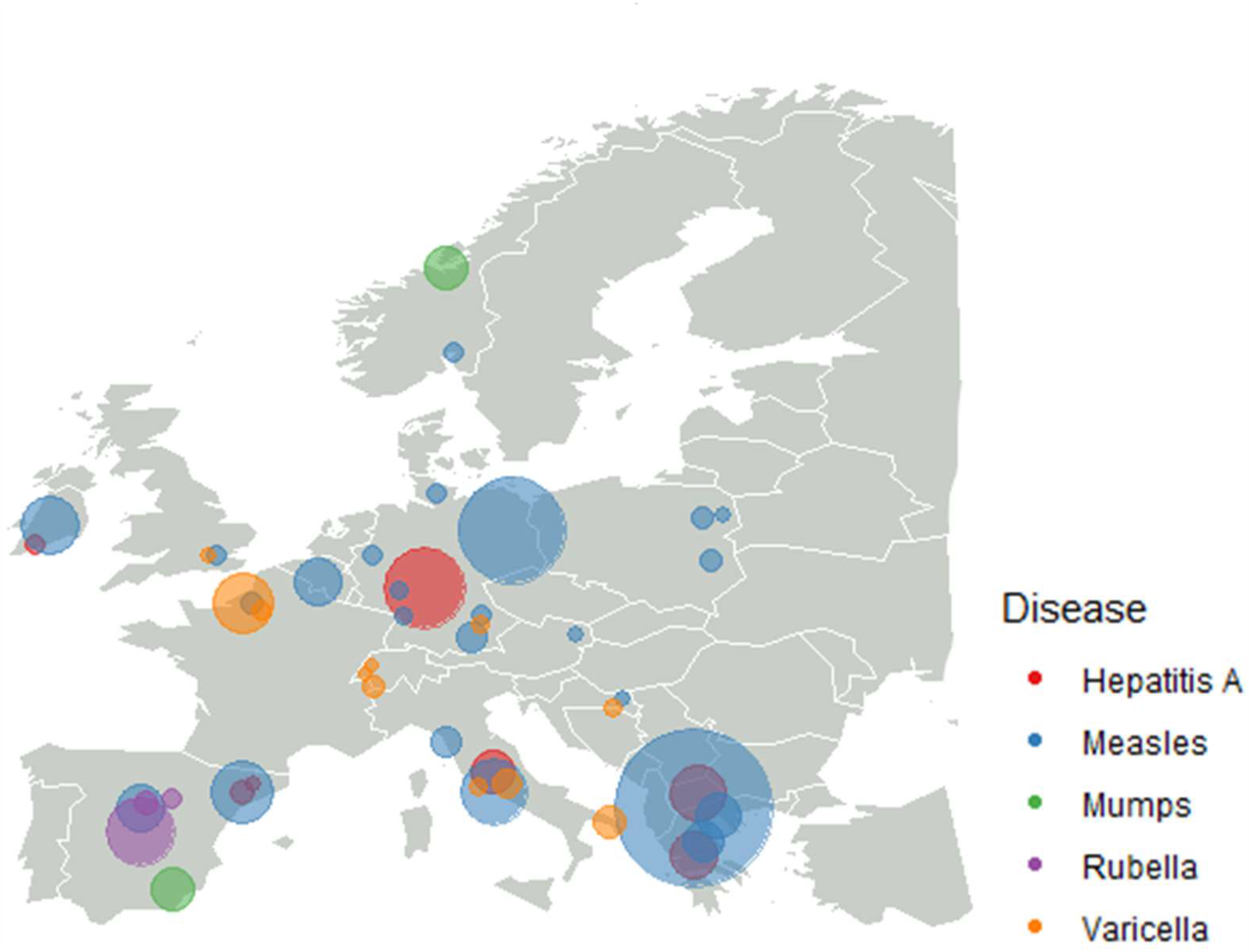
Case-load of vaccine-preventable disease outbreaks involving migrants by reported location. Bubble colours represent disease type, bubble size represents caseload.

### Measles outbreaks in migrant populations

21 reports discussing measles outbreaks were included, reporting on 24 distinct outbreaks and one report that presented all outbreak-related cases in asylum seekers in Germany from 2004 to 2014. Seven reports originated from Germany (16, 24, 29, 37, 52, 56, 59), four from Greece (50, 51, 54, 55), three from Italy (46, 47, 58), two from Spain (45, 48) and one each from Croatia (53), France (26), Ireland (49), Norway (57) and Poland (35). A large proportion of outbreaks were associated with Eastern European migrants, particularly of Roma nationality, including nine outbreaks with an Eastern European index case (16, 37, 46, 47, 51, 52, 58, 62, 67, 68). Measles outbreaks were frequently observed in refugee camps, residences or shelters, with a high burden in children and adolescents in both Roma communities and refugee camps.

Between August 2009 and July 2011, a Europe-wide outbreak occurred, which peaked in March 2010 with 6,664 cases (14) and arose from a new strain of measles virus, D4-Hamburg, which notably led to over 24,000 cases in Bulgaria (62). Nine reports of measles outbreaks involving migrants were included from this time period (29, 49, 52-54, 56, 57, 62), of which six had a high burden in migrants from Eastern European countries, mostly of Roma ethnicity (49, 52, 54-56, 62) and four had a Bulgarian index case (52, 54-56, 62). These outbreaks were often linked to very low vaccination coverage in the affected populations. In one Greek outbreak, none of the migrant cases had a recorded measles vaccination and in an Irish outbreak, only 14% and 2% of cases had evidence of having had one and two measles vaccines, respectively (49, 54). The Greek outbreak led to 83 hospitalisations, whereas in Ireland 115 hospitalisations occurred.

Post-2015, which saw a major influx of migrants to Europe (69), four measles outbreaks involving migrants were reported, two of which took place in hosting facilities for asylum seekers and refugees (16, 26). Another took place in Greece across 2017 and 2018 and was responsible for 3150 cases, of whom the majority (80.4%) were unvaccinated, with three Romanian Roma siblings reported as the index cases (51). Over half (61.3%) of the cases were hospitalised, with complications reported in 17.1% of cases and four deaths.

### Varicella in migrant populations

Ten reports discussing varicella outbreaks were included, reporting on eleven distinct outbreaks and one report that presented all outbreak-related cases in asylum seekers in Germany from 2004 to 2014. Three reports came from Italy (33, 36, 60), two from France (27, 30) and Germany (24, 32) and one each from Croatia (53), England (66) and Switzerland (31). Ten of the included varicella outbreaks (83.3%) took place in refugee camps or asylum centres, of which only one case resulted in spread to the general population; the son of an asylum centre worker was infected following an outbreak in the centre (33). Populations involved in varicella outbreaks were mainly African, Middle Eastern or South Asian, with the most frequently-reported nationalities involved being Sudanese (27, 30, 33) and Somalian (31, 32, 66) (both n=3) followed by Eritrean (27, 31) and Sri Lankan (31, 33) (both n=2). In all five papers where age-groups involved were reported, the populations affected were adults.

Following the major influx of migration to Europe in 2015, a varicella outbreak took place in an asylum centre in Italy, infecting 41 individuals of mostly East and West African origin (36). Following this outbreak, serology testing was done in the asylum centre, showing that >20% of East and Central Africans and South Asians were susceptible. Two outbreaks affected refugee camps in the Calais area, with the first resulting in 12 cases, of whom two were hospitalised for respiratory involvement, and the second 351 cases, of whom 90% were over 15 years old and two went on to suffer from pulmonary complications (27, 30).

### Hepatitis A outbreaks in migrants

Eight papers included in this review reported on hepatitis A outbreaks involving migrants. Two papers reported on outbreaks occurring in Germany (24, 34), two in Spain (25, 43), two in Greece (28, 42) and one each in Ireland (41) and Italy (40). A total of 1,510 cases of hepatitis A were reported as part of outbreaks involving migrants, of which 1,322 (87.5%) occurred after January 2015. Outbreaks affecting migrants were predominantly reported to be of sub-genotype IB, similar to strains previously isolated from samples in the Middle East, Turkey, Pakistan and East Africa (28, 34, 42), in contrast to recent European hepatitis A outbreaks in native populations, where sub-genotype IA is often prevalent (34, 70-72).

Between September 2015 and March 2016, cases rose across Germany by 45% from what is to be expected (34). This dramatic increase was due to an unusually high burden of disease in migrants; of the 699 cases, 278 (40%) were in asylum seekers, 90% of which were of sub-genotype IB. In 2016 and 2017, Greece reported 188 and 30 cases, respectively, of hepatitis A of mainly sub-genotype IB in the childhood refugee population (28). The individuals affected were mainly Syrian, with some Iraqi and Afghan nationals also reported as positive.

### Rubella outbreaks in migrants

Three rubella outbreaks involving migrants were reported, all occurring in Spain (44, 61, 64, 65). Overall, 487 cases of rubella were included, of which 277 (56.9%) were migrants. Across all outbreaks, the migrants affected were predominantly Latin American, with two affecting only this group (61, 64). Around a third of cases (151; 31.0%) were reported as being in women of child-bearing age, of whom seven were pregnant and subsequently terminated their pregnancies. In two outbreaks, all affected individuals reported being unvaccinated. In the remaining outbreak, 94.3% (n=460) did not have evidence of rubella vaccination (65).

### Mumps and other VPDs in migrants

Mumps outbreaks were reported in three studies (295 cases), including two distinct outbreaks (39, 63) and one report that presented all outbreak-related cases in asylum seekers in Germany from 2004 to 2014 (24). One outbreak in Spain across 17 municipalities of Almeria (63) caused 145 infections, of which a small number (4 cases) were migrants. Another outbreak in Norway was reported as having a foreign student as the index case and went on to infect 148 individuals, resulting in seven hospitalisations and one case of meningitis (39).

## Discussion

Our data highlight that migrants are an at-risk group involved in VPD outbreaks in Europe. We have shown that adult and child migrants living in temporary shelters or camps are at particular risk, alongside specific nationality groups. Measles, varicella and hepatitis A were responsible for the largest burden of outbreak-related VPDs in migrants. A large majority of included varicella outbreaks were associated with adults, whereas measles and hepatitis outbreaks affected a wide range of age-groups. Half of included measles outbreaks were associated with migrants from Eastern European countries, often of Roma ethnicity, with several outbreaks leading to high numbers of hospitalisations, complications and deaths. Our data highlight the importance of tailoring strategies for implementing catch-up vaccination to specific at-risk groups, alongside the strengthening of routine data collection, in order to avoid outbreaks of vaccine-preventable disease.

Measles, varicella and hepatitis A outbreaks were frequently reported from hosting facilities for refugees and asylum seekers, making up almost half of all outbreaks included in this systematic review. In these outbreaks, secondary cases among the local population were mostly limited, but large numbers of migrants were often affected, for example, 351 migrants in a camp in Calais during an outbreak of varicella (30). Since the large influx of migration in 2015, temporary camps have become a new feature in Europe and the numbers of forced migrants residing in such conditions continues to increase, with Greece alone hosting almost 40,000 migrants living in temporary reception centres in 2020 (73). Many stay for extensive time periods in these settlements, which often run well beyond capacity and in suboptimal conditions, including lack of basic infrastructure and sanitation facilities. In most cases, migrants residing in temporary camps are not included in national vaccination systems, which could mean children miss their routine vaccinations. Poor sanitation makes these settlements highly conducive to outbreaks such as hepatitis A, which can be spread through poor sanitation (5, 42). Despite this, limited interventions exist in European countries to provide vaccination services to those living in refugee camps, with NGOs often left to fill the gap (74).

Hepatitis A outbreaks among migrants were often associated with living conditions and genetically unrelated to outbreaks in the general population, amongst whom MSM (men who have sex with men) can be considered the main risk group (42). Varicella, which we have shown often causes outbreaks amongst adults in temporary camps or detention centres, some of whom required hospitalisation, is associated with more severe outcomes in adults (75). Whilst most children born in Europe will have varicella at a young age and gain immunity, less than 60% of adults from tropical countries have immunity through a varicella episode during childhood (75, 76). Varicella is not part of routine vaccination schedules in most European countries; however, our data suggests that the impact on communities living in refugee camps justifies the implementation of organised vaccination in these settings. Provision of sanitary living conditions and appropriate healthcare in refugee camps and detention centres, alongside universal vaccination of residents for key diseases, such as measles, varicella and hepatitis A, are essential to avoid further mortality. In 2014, a measles outbreak amongst young asylum seekers spread to the wider population, causing a total of 1,344 cases and leading to 345 hospitalisations and the death of a 1-year old child (37), demonstrating the potential consequences of neglecting at-risk populations.

A key risk-factor for outbreaks is under-immunisation in the affected populations, with affected migrant groups often having particularly low vaccine coverage. Numerous previous studies have suggested migrants are an under-immunised group; we have shown in a previous systematic review and meta-analysis that migrants represent an under-immunised population in the EU for measles, mumps, diphtheria and tetanus (77). Another review has also suggested low immunity coverage in migrants and refugees to the EU compared to the host population (2). A recent study reported that the only factor associated with increased measles incidence is low vaccine coverage, rather than migration itself; the study highlights the current low coverage of the second vaccine dose (MCV2) in many Eastern European countries, in particular Romania (coverage of 75%), a country which also has the highest measles incidence rate in the region (46.1/100,000) (78). This reflects our findings that movement of under-immunised children from Eastern Europe, mainly of Roma ethnicity, combined with pockets of under-immunisation in host countries are a linking factor between outbreaks of measles. Roma communities are an under-immunised group in a variety of settings in Europe and face many barriers to accessing vaccination, including language, trust in health services, and concerns around vaccine safety (79-82). Robust scientific messaging to address community-specific trust issues and circulating misinformation has been highlighted as a key component of COVID-19 vaccine roll-out (11, 12). It is crucial going forward that interventions are developed and tailored towards at-risk migrant communities to build trust and reduce disparities in access to vaccination systems (83). Migrants should have access to key vaccines regardless of their immigration status: the IOM has specifically called for governments to ensure an adequate COVID-19 vaccine stock be reserved for non-nationals and forcibly displaced migrants, regardless of their immigrations status (84).

Whilst our systematic review has brought to light several key at-risk populations groups for specific diseases in Europe, the scope of the results is limited by the availability and quality of the datasets that have been published. Despite including ten distinct VPDs in our search strategy, only five diseases (measles, varicella, hepatitis A, rubella and mumps) returned any reports that met our inclusion criteria. This may be due to a higher outbreak-causing potential of these diseases compared to, for example, diphtheria, where isolated cases in migrants were often reported but did not appear to spread (85-87). Few studies compare migrant incidence or involvement in outbreaks with non-migrants, making it difficult to put these data into context and determine which groups are most vulnerable. Differences in reporting systems between European countries is inhibitory to the assimilation of accurate data on at-risk groups for VPD outbreaks. It is of utmost importance that demographical data on outbreaks (including nationality and migrant status) is collected and reported in a transparent and disaggregated manner, for example, as part of the European Surveillance System (TESSy), as this can provide insight into populations who may be at particular risk as well as health system or technological weaknesses that may leave an area or population open to future outbreaks.

Recent ECDC guidelines (88) call on EU/EEA countries to offer vaccination against measles, mumps and rubella (MMR) to all migrant children and adolescents as well as adults without immunisation records, in accordance with the immunisation schedule of the host country. Adult migrants should also be given a primary series of diphtheria, tetanus, and polio vaccines. We have also shown in this systematic review the risks associated with lack of immunity to varicella and hepatitis A, particularly in refugee camps, suggesting specific guidelines should also be developed to cover these vaccinations for migrants. These data provide a clear rationale for greater emphasis to be placed on developing effective and targeted strategies to improve vaccine coverage in migrants as an at-risk group, particularly for adolescent or adult migrants who will not be easily incorporated into host vaccination schedules on arrival. This will require further research to better understand vaccine uptake and demand issues in migrant groups (including confidence, convenience and complacency) and greater focus on co-designing vaccine uptake strategies in close collaboration with affected communities, as well as commissioning of health services and offering incentives for intervention development. Ensuring clinicians have access to appropriate guidelines and heightened awareness of potential under-immunisation in at-risk groups presenting is a vital next step in improving vaccine coverage and preventing outbreaks and cases of preventable VPDs going forward. A specific focus needs to be placed on catch-up vaccination for adults who may be excluded from routine immunisation schedules in their host country, with an emphasis on community engagement to ensure acceptability. It is also crucial that vaccination efforts in refugee camps, where poor living conditions leave many at risk, are prioritised using NGOs or other groups who have already developed connections and trust in these settings.

## Data Availability

All raw data is available on request

## Conflicts of interest

All authors report having nothing to declare

## Contributions

SH had the idea for this review. AD and SH designed the protocol. AD led the searches, data extraction/analysis, with input from RH and SEH. AD and SH wrote a first draft of the paper. All authors viewed and discussed the data and contributed to the writing of the paper.

## Acknowledgements

This work has been funded by the NIHR (NIHR300072). AD and SEH are funded by the MRC (MR/N013638/1). SH is funded by the NIHR (NIHR Advanced Fellowship NIHR300072) and the Academy of Medical Sciences (SBF005\1111). This work has been supported by the European Society of Clinical Microbiology and Infectious Diseases (ESCMID) Study Group for Infections in Travellers and Migrants (ESGITM). Kieran Rustage is funded by the Rosetrees Trust (M775). SMJ was funded by the National Institute for Health Research Health Protection Research Unit (NIHR HPRU) in Immunisation at the London School of Hygiene and Tropical Medicine (LSHTM) in partnership with Public Health England (PHE).

## Supplementary information: Key messages

### Panel: Key messages

Migrants are an at-risk group involved in VPD outbreaks in Europe and comprise one of several under-immunised groups in the Region.

Adult and child migrants living in temporary shelters, camps or detention centres are at high risk from VPD outbreaks, particularly of measles, varicella and hepatitis A.

At-risk groups varied by disease; a large majority of varicella outbreaks were associated with adult migrants, whereas half of included measles outbreaks were associated with migrants of all ages from Eastern European countries, often of Roma ethnicity.

Our data highlight the importance of tailoring vaccine-delivery strategies to specific at-risk migrant groups, and strengthening routine data systems to capture data on uptake and coverage, in order to meet regional and global vaccination targets and avoid outbreaks.

Migrants may face barriers to vaccination on arrival, including exclusion from vaccination systems, with important implications for COVID-19 vaccine delivery going forward. A better understanding of vaccine uptake and demand issues in migrant groups is urgently needed, alongside a greater focus on co-designing vaccine uptake strategies in close collaboration with affected migrant communities.

## Supplementary Information: Search Strategy

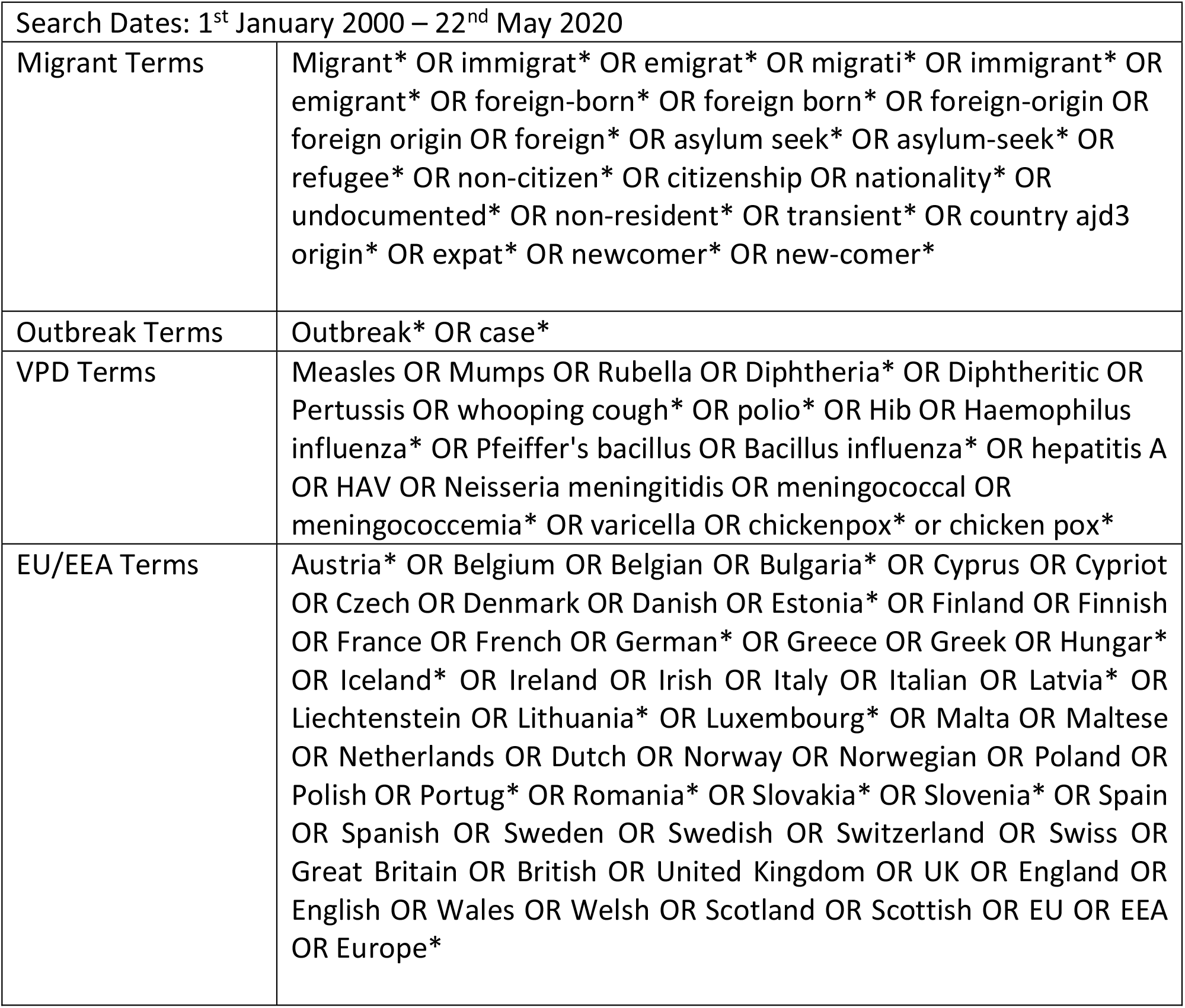

## Notes

### Competing Interest Statement

The authors have declared no competing interest.

### Clinical Protocols

https://www.crd.york.ac.uk/PROSPERO/display_record.php?RecordID=157473

### Author Declarations

As a systematic review, this study is exempt from IRB/ethical approval.

